# Computational Decomposition of New Memory Failure in Alzheimers Disease Through a Hippocampal Cortical Consolidation Bottleneck Model

**DOI:** 10.64898/2026.06.23.26356309

**Authors:** Meiwei Zhang, Yuwei Pan, Lihua Chen

**Affiliations:** College of Electrical Engineering, Chongqing University, Chongqing, 400030, China; Department of Geriatrics, The First Affiliated Hospital of Chongqing Medical University, Chongqing, 400016, China

**Keywords:** Alzheimers disease, Memory consolidation, Computational phenotyping, Neurodynamic rigidity

## Abstract

**Objective:** Alzheimers disease (AD) is characterised by difficulty retaining newly learned information, but routine memory scores often conflate poor initial encoding with impaired post-encoding stabilisation. This study aimed to develop an interpretable computational phenotype that separates new-memory failure during progression from mild cognitive impairment (MCI) to AD.

**Methods:** We proposed a Hippocampal–Cortical Consolidation Bottleneck (HCCB) model, representing newly learned information as a rapidly formed hippocampal trace and a slowly stabilised cortical trace. The model predicts a residual bottleneck when delayed recall is lower than expected from immediate recall. This prediction was operationalised as the Consolidation Bottleneck Index^∗^ (CBI^∗^), a cognitively normal reference-normalised residual index. CBI^∗^ was evaluated in ADNI participants spanning normal cognition, MCI nonconversion, MCI conversion and AD, using cognitive and MRI data. Independent neurodynamic support was examined using OpenNeuro resting-state EEG.

**Results:** Simulations showed new-memory vulnerability when hippocampal vulnerability exceeded cortical vulnerability. In ADNI, CBI^∗^ increased across the clinical spectrum and reached AD-like levels in MCI converters. Higher CBI^∗^ was associated with hippocampal atrophy, supporting its anatomical relevance. CBI^∗^ added limited discrimination beyond established clinical and structural predictors, indicating that it captured a mechanistic phenotype rather than serving as a replacement prognostic model. OpenNeuro EEG further showed increased neurodynamic rigidity in AD.

**Conclusions:** The HCCB framework quantifies failed stabilisation of newly encoded information and links this phenotype to hippocampal degeneration and altered neurodynamics.

**Significance:** This study provides an interpretable computational framework for characterising consolidation failure in AD progression.

## 1. Introduction

Alzheimers disease (AD) is one of the leading causes of cognitive decline in older adults and is clinically characterised by progressive impairment in memory, cognition and daily function (McKhann et al., 2011; Jack et al., 2018; Zhang et al., 2024; Parameswari et al., 2026). Among its most recognisable clinical features is the difficulty of retaining newly learned information, while more remote memories may be relatively preserved during earlier disease stages. This temporal pattern has long been discussed in relation to hippocampal vulnerability, system consolidation and the staged spread of AD pathology (Braak and Braak, 1991; Squire and Alvarez, 1995; Nadel and Moscovitch, 1997; Frankland and Bontempi, 2005). The complementary learning systems theory further provides a computational account of why rapid hippocampal encoding and slower neocortical learning may support different stages of memory formation (McClelland et al., 1995; O’Reilly et al., 2014). However, the clinical familiarity of this phenomenon does not resolve a more basic computational problem. Routine memory assessment usually reports how much a person recalls, but it does not specify which component of the memory process has failed.

A low delayed recall score can arise from at least two different mechanisms. One patient may perform poorly because the information was not encoded effectively at the outset. Another may encode the information reasonably well, but fail to stabilise it over time (Marvi et al., 2024). The distinction between initial encoding and later consolidation is central to neurobiological theories of memory, in which a newly formed trace may remain labile before becoming stabilised through hippocampal cortical interactions (Dudai, 2004; Winocur et al., 2010). These two cases can appear similar if delayed recall is considered alone, yet they imply different failure modes. This distinction is especially important for computational phenotyping, because a clinically useful model should not only measure that memory is impaired, but also clarify whether the impairment reflects poor initial learning or failed post encoding stabilisation (Zhao et al., 2025; Boccalini et al., 2025). This motivates a hippocampal cortical formulation in which newly presented information first forms a fast but fragile hippocampal trace and is then progressively stabilised into slower cortical representations.

This decomposition also raises a practical measurement question. If failed stabilisation is clinically meaningful, it should be observable in routine cohort data without requiring specialised autobiographical memory tasks (Donarelli et al., 2025). Delayed recall tests are widely used in AD assessment and have been proposed as clinically meaningful markers of AD-related memory impairment (Cerami et al., 2017a; Tong et al., 2016). The Alzheimer’s Disease Neuroimaging Initiative (ADNI) provides a large longitudinal resource for connecting such neuropsychological measures with diagnosis, genotype, imaging biomarkers and clinical progression (Mueller et al., 2005; Petersen et al., 2010a; Veitch et al., 2024). ADNI-MEM has also been developed as a composite memory measure integrating several ADNI memory tasks (Crane et al., 2012). However, composite memory scores and delayed recall scores usually quantify overall memory impairment rather than the residual loss of delayed recall after accounting for initial encoding. Delayed recall should therefore be interpreted relative to immediate recall, rather than as an isolated score (Russo et al., 2017). A person whose delayed recall is low because immediate recall was already poor should not be treated the same as a person whose delayed recall falls below what would be expected from their initial encoding level. This motivates a residual bottleneck phenotype, Consolidation Bottleneck Index^∗^ (CBI^∗^), that quantifies delayed recall loss beyond the level predicted by immediate recall and demographic factors in a cognitively normal reference model. Such a phenotype can be tested in ADNI to determine whether future mild cognitive impairment converters already show an Alzheimer like pattern of post encoding stabilisation failure.

A further question is whether this residual bottleneck reflects a biologically meaningful process rather than a statistical transformation of cognitive scores. Hippocampal atrophy has been repeatedly associated with AD progression and conversion from mild cognitive impairment to AD (Jack et al., 1999; Apostolova et al., 2006; Jack et al., 2013). If the bottleneck captures failed stabilisation of newly encoded information, it should therefore be related to medial temporal degeneration, particularly hippocampal atrophy. At the same time, stabilisation of new information is unlikely to depend only on static anatomy. It also requires a brain state capable of flexible reconfiguration, sustained information processing and dynamic response to input. Resting EEG studies have reported AD-related slowing, altered synchrony and reduced signal complexity (Jeong, 2004; Dauwels et al., 2010; Cassani et al., 2018). Recent public EEG datasets further enable reproducible analyses of dementia-related electrophysiological alterations across AD, frontotemporal dementia and healthy controls (Miltiadous et al., 2023). This motivates a complementary electrophysiological analysis using OpenNeuro EEG data to test whether Alzheimer’s disease is accompanied by reduced neurodynamic flexibility. EEG is not used here as a direct measure of memory consolidation (Kanda et al., 2017), but as an independent test of the model’s dynamical assumption that Alzheimer’s disease occurs in a more rigid neural regime.

The overall study design is summarised in Figure 1. Starting from the ambiguity between poor initial encoding and failed post encoding stabilisation, the proposed Hippocampal Cortical Consolidation Bottleneck (HCCB) model derives a residual consolidation bottleneck phenotype, which is then evaluated using ADNI cognitive and MRI data and independently supported by OpenNeuro EEG evidence of altered neurodynamic flexibility.

**Figure 1:**
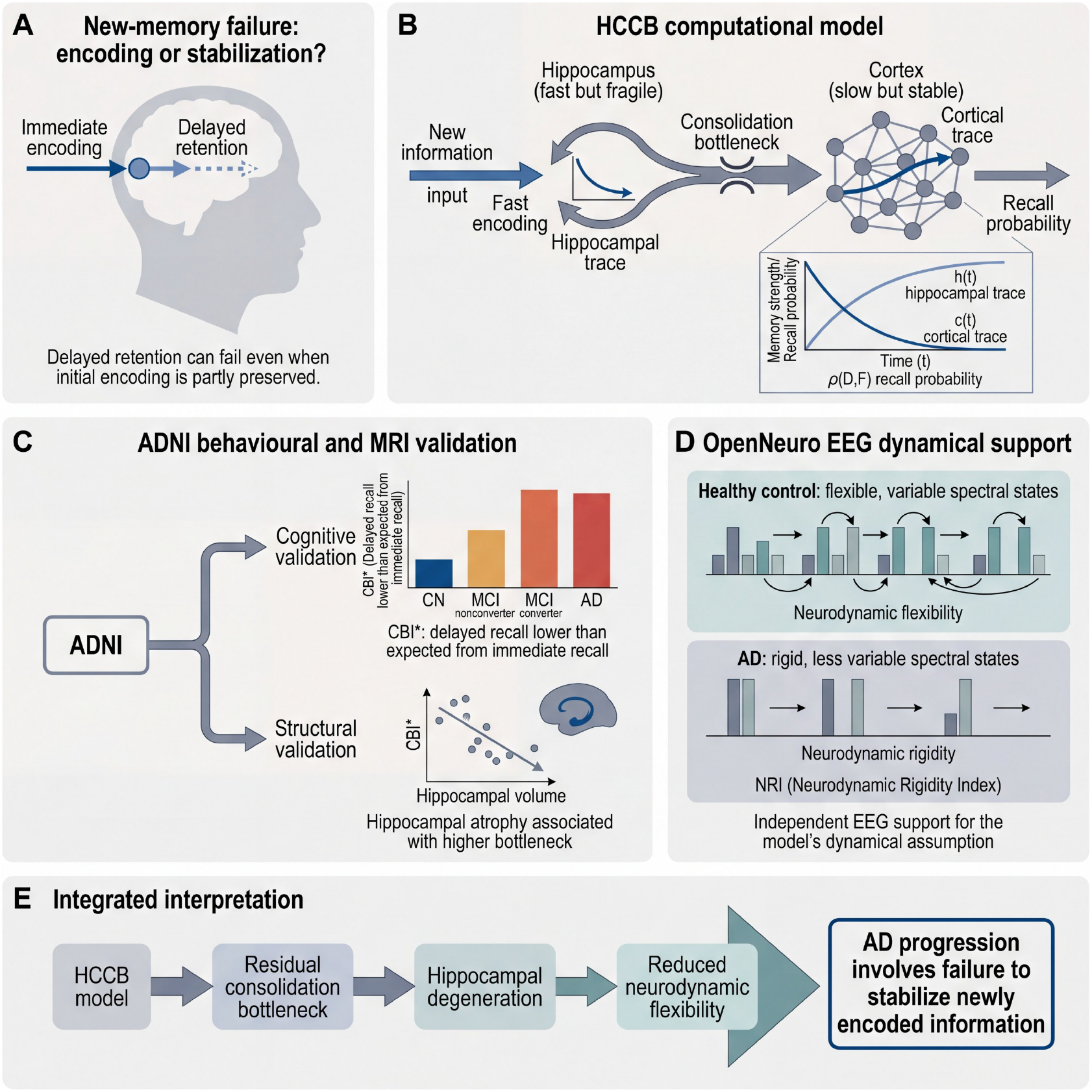
Overview of the proposed HCCB framework and validation strategy. The study starts from the clinical ambiguity between poor initial encoding and failed delayed retention, formalises this distinction using a hippocampal cortical consolidation bottleneck model, validates the residual bottleneck phenotype using ADNI cognitive and MRI data, and uses OpenNeuro EEG to provide independent support for reduced neurodynamic flexibility in Alzheimer’s disease.

The contributions of this paper are summarized as follows:

- A computational decomposition model, termed Hippocampal Cortical Consolidation Bottleneck, is developed to separate poor initial encoding from failed post encoding stabilisation in new memory failure.
- A measurable residual bottleneck phenotype, CBI^∗^, is formulated from routine clinical memory testing to quantify delayed recall loss not explained by immediate recall or demographic factors. In ADNI, this phenotype identifies an Alzheimer like stabilisation failure pattern in mild cognitive impairment converters.
- A cross level validation strategy is established by linking CBI^∗^ to hippocampal atrophy in ADNI and by testing the model’s neurodynamic assumption using OpenNeuro EEG evidence of reduced flexibility in Alzheimers disease.

The rest of this article is organized as follows. Section II reviews related work. Section III presents the proposed HCCB-based computational framework. Section IV describes the experimental setup. Section V reports the results and discussion. Section VI concludes the paper and outlines future work.

### 2. Related work

Computational theories of memory consolidation have long emphasised the complementary roles of hippocampus and neocortex. The complementary learning systems framework argues that the hippocampal system supports rapid encoding of new experiences, whereas the neocortex gradually extracts structured knowledge across repeated experiences (McClelland et al., 1995; O’Reilly et al., 2014). Standard systems consolidation theory further suggests that memories can become progressively reorganised from hippocampal dependent representations toward more distributed cortical representations (Squire and Alvarez, 1995; Winocur et al., 2010). These theories provide a strong neurocomputational basis for understanding why newly encoded information is vulnerable in Alzheimer’s disease. However, most existing frameworks remain conceptual with respect to routine clinical memory testing. They do not directly specify how to separate poor initial encoding from failed post encoding stabilisation in cohort scale clinical data. HCCB builds on this literature by converting the hippocampal cortical consolidation idea into a dynamical model that explicitly distinguishes the fast encoding trace from the slower stabilisation trace.

Clinical studies have consistently shown that delayed recall is sensitive to Alzheimer’s disease and disease progression. Delayed recall has been discussed as a candidate gateway biomarker for Alzheimer’s disease, and ADNI has provided a major longitudinal resource for connecting neuropsychological measures, MRI, genetics and clinical progression (Cerami et al., 2017b; Petersen et al., 2010a). ADNI-MEM further summarises episodic memory performance across multiple memory tasks, including RAVLT, ADAS-Cog, MMSE memory items and Logical Memory, and has been widely used as a composite memory outcome (Crane et al., 2012). Structural MRI studies have also shown that hippocampal atrophy predicts conversion from mild cognitive impairment to Alzheimer’s disease (Jack et al., 1999; Apostolova et al., 2006). These studies establish the importance of memory and medial temporal degeneration, but they mainly treat memory decline as an overall cognitive phenotype or prognostic marker. In contrast, the present study focuses on a residual stabilisation phenotype. CBI^∗^ asks whether delayed recall is lower than expected from immediate recall and demographic factors, thereby isolating the component of memory failure that is more consistent with failed post encoding stabilisation.

EEG has been widely studied as a low cost and temporally precise modality for Alzheimer’s disease assessment. Prior work has reported characteristic EEG slowing, reduced signal complexity and altered synchrony in Alzheimer’s disease (Jeong, 2004; Dauwels et al., 2010). Systematic reviews of resting state EEG have further summarised its potential for Alzheimer’s disease diagnosis and progression assessment, while also noting variability in preprocessing, feature extraction and validation strategies across studies (Cassani et al., 2018). The OpenNeuro ds004504 dataset provides resting state EEG recordings from Alzheimer’s disease, frontotemporal dementia and healthy control participants, enabling reproducible analysis of dementia related EEG alterations (Miltiadous et al., 2023). Most EEG studies, however, use EEG as a diagnostic or classification signal. In this study, EEG is used differently: it tests a dynamical assumption of HCCB. Specifically, neurodynamic rigidity is treated as an electrophysiological signature of reduced flexibility, which may constrain the stabilisation of newly encoded information. This positions EEG as independent support for the model’s dynamical component rather than as direct evidence of memory consolidation.

#### Algorithm 1

HCCB-based computation and validation pipeline

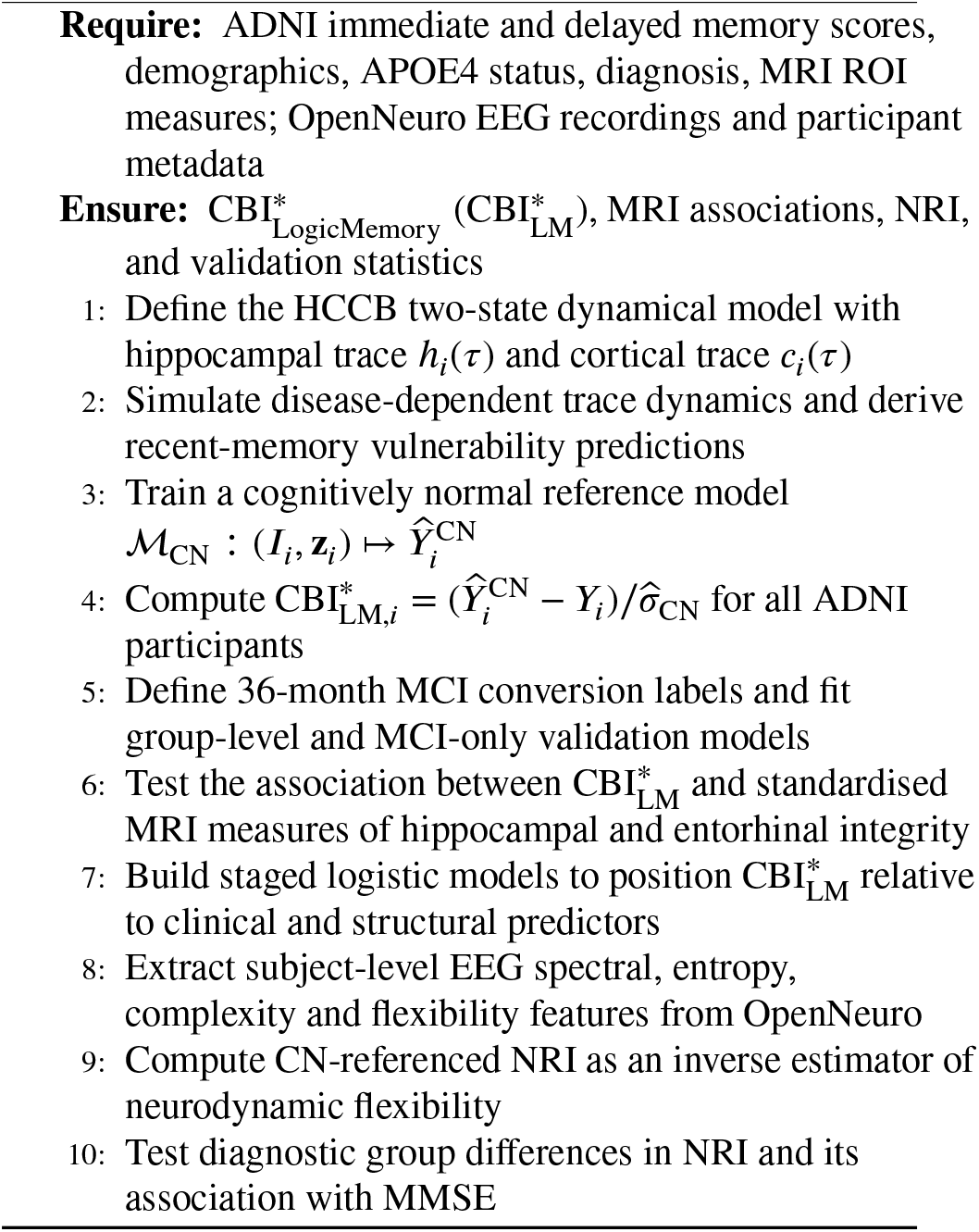

## 3. Proposed Method

The proposed framework consists of four computational components: a hippocampal cortical state-space model, a normative residual estimator for clinical memory data, a clinical and structural validation protocol, and an EEG-based rigidity estimator. The dynamical model is used to derive observable quantities rather than to fit dense individual memory trajectories, which are not available in ADNI (Petersen et al., 2010b). The resulting pipeline is summarised in Algorithm 1.

### 3.1. HCCB State-Space Model

The Hippocampal Cortical Consolidation Bottleneck model was developed to separate two components of new-memory failure that are usually mixed in delayed recall scores. A low delayed recall score may indicate poor initial encoding, failed stabilisation after encoding, or both. HCCB represents this distinction as a two-state nonlinear dynamical (Kaplan and Glass, 2012; Yu et al., 2025) system:

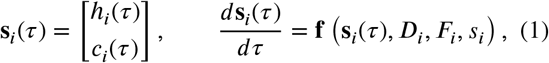

where *h*_*i*_(*τ*) is a fast hippocampal trace, *c*_*i*_(*τ*) is a slower cortical trace, *D*_*i*_ denotes disease load, *F*_*i*_ denotes neurodynamic flexibility, and *s*_*i*_ denotes schema support. The model was used as a generative analytical model to derive empirical estimators and validation targets, rather than as a full participant-wise fit to ADNI or EEG trajectories.

For a newly presented memory item, the initial state is

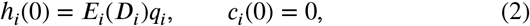

where *q*_*i*_ denotes item-level encoding quality. Disease modulates the effective encoding capacity as

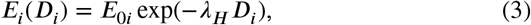

where *λ*_*H*_ controls the disease sensitivity of the hippocampal encoding component (Daou et al., 2023).

After encoding, the hippocampal trace decays and contributes to cortical stabilisation. The coupled dynamics are

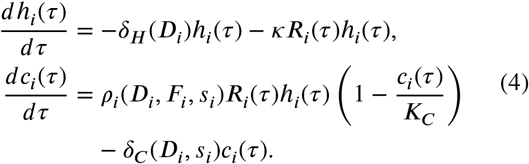

Here, *R*_*i*_(*τ*) denotes reactivation strength, *κ* controls hippocampal-to-cortical transfer, and *K*_*C*_ is the effective cortical storage capacity. The consolidation rate is parameterised as

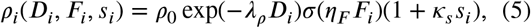

where *σ*(*z*) = 1/(1 + exp(−*z*)). This form encodes three assumptions: disease load reduces stabilisation, neurodynamic flexibility facilitates stabilisation, and schema support improves integration into existing cortical structure. The cortical decay term is

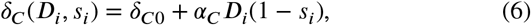

so that schema-supported traces are less vulnerable to disease-related cortical destabilisation.

Recall probability is generated from the state vector through a readout function:

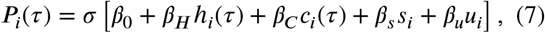

where *u*_*i*_ denotes retrieval support. The simulated Consolidation Bottleneck Index between an immediate time point *τ*_imm_ and a delayed time point *τ*_del_ is

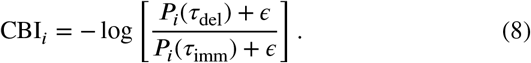

A larger value indicates greater loss of delayed recall relative to immediate recall.

The model also yields a simple recent-memory vulnerability condition (Park and Kaang, 2026). Let

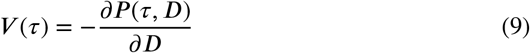

denote disease sensitivity at memory age *τ*. When the hippocampal component is more disease-sensitive than the cortical component, the model predicts a recent-memory vulnerability regime:

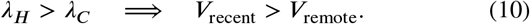

This condition was examined numerically across the (*λ*_*H*_, *λ*_*C*_) parameter space. To model remote memories more realistically, a two-stage simulation was used. A memory was first consolidated under *D* = 0 for a specified pre-disease interval. Disease load was then introduced, and post-onset loss was computed as

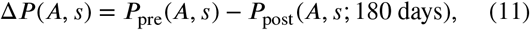

where *A* is the memory age at disease onset and *s* is schema support.

### 3.2. CBI* Estimation From ADNI

ADNI was used to estimate whether the model-derived bottleneck can be measured from routine clinical memory data. Diagnostic, demographic, genetic, neuropsychological and MRI-derived variables were harmonised at the participant level using *RID*. The primary clinical groups were cognitively normal participants, mild cognitive impairment nonconverters, mild cognitive impairment converters and Alzheimer’s disease patients. For the 36-month conversion analysis, baseline MCI participants who converted to AD within 36 months were labelled converters. Baseline MCI participants who did not convert within 36 months were labelled nonconverters for that prediction window. Participants with insufficient follow-up and no observed conversion were excluded from the fixed-window conversion analysis.

Logical Memory was used as the primary cognitive measure because it contains both immediate and delayed recall (Clarke et al., 2022). Let *I*_*i*_ denote Logical Memory immediate recall and *Y*_*i*_ denote Logical Memory delayed recall (Zheng et al., 2023). A raw bottleneck score was first defined as

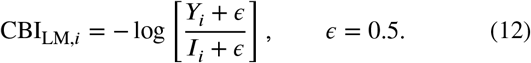

This ratio is interpretable but remains sensitive to baseline learning level and demographic effects. The primary estimator therefore used a cognitively normal reference model.

Let ℳ_CN_ denote a normative delayed-recall estimator trained only in cognitively normal participants:

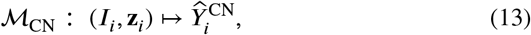

where **z**_*i*_ includes age, sex and education. In this study, ℳ_CN_ was implemented as a linear reference model:

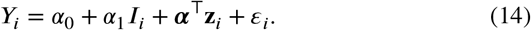

The fitted model was then applied to all participants:

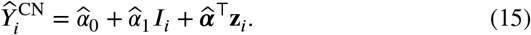

The residual bottleneck estimator was defined as

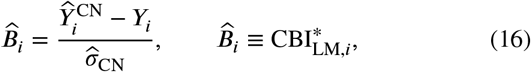

where 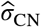 is the standard deviation of residuals in the cognitively normal reference group. A higher value indicates that delayed recall is lower than expected from immediate recall and demographic factors. In the HCCB interpretation, this residual is an observable estimator of post-encoding stabilisation failure.

RAVLT was analysed as a secondary memory measure. To keep the immediate and delayed measures on the same scale, trial 5 was used as the immediate reference:

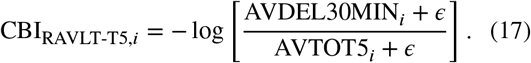

Structural MRI analysis used intracranial volume-adjusted hippocampal volume,

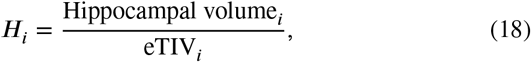

and entorhinal cortical thickness. MRI predictors were standardised before regression analysis:

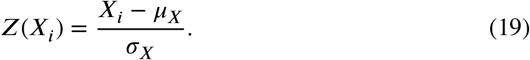

### 3.3. Clinical and Structural Validation

The first validation test examined whether 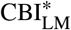 increased across the AD clinical spectrum. The adjusted^L^g^M^roup model was

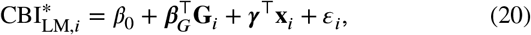

where **G**_*i*_ encoded diagnostic group and **x**_*i*_ included age, sex, education and APOE4 dose. Cognitively normal participants served as the reference group. HC3 robust standard errors were used. Planned contrasts compared MCI converters with MCI nonconverters, AD with MCI nonconverters and MCI converters with AD.

To test the prodromal stage directly, a second model was fitted only in baseline MCI participants:

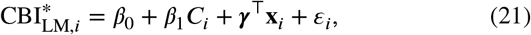

where *C*_*i*_ = 1 denotes 36-month conversion from MCI to AD and *C*_*i*_ = 0 denotes nonconversion within the same window. This model tests whether future converters already show an elevated residual bottleneck while they are still clinically classified as MCI.

MRI validation examined whether the residual bottleneck was structurally anchored to medial temporal degeneration:

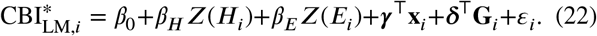

Here, *H*_*i*_ is ICV-adjusted hippocampal volume and *E*_*i*_ is entorhinal thickness. Hippocampus-only, entorhinal-only and joint MRI models were estimated in the full sample. A corresponding MCI-only MRI model was also fitted to evaluate whether the structural association was already present during the prodromal stage.

Finally, staged logistic models were used for prognostic positioning. The purpose was not to claim a new optimal conversion classifier, but to determine whether 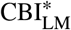 provided information beyond established clinical and str^L^u^M^ctural markers. The outcome was 36-month MCI-to-AD conversion:

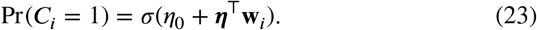

The base model included age, sex, education and APOE4 dose. The clinical model added MMSE, ADNI-MEM and CDR-SB. The structural model added ICV-adjusted hippocampal volume. The full model added 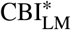. Model discrimination was summarised by AUC:

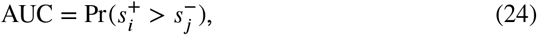

where 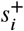 and 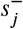 are predicted risks for converters and nonconverters. DeLong testing was used to compare the structural model with the full model. Sensitivity analyses examined outlier trimming, winsorisation, visit-date restrictions, alternative conversion windows and exclusion of very low immediate recall values.

### 3.4. EEG Rigidity Estimation

OpenNeuro EEG data were used to evaluate the dynamical assumption of HCCB that AD is accompanied by reduced neurodynamic flexibility. The resting-state dataset ds004504 was used as the primary EEG analysis (Markiewicz et al., 2021). The photic-stimulation dataset ds006036 was analysed as an exploratory test of stimulus-driven adaptability. EEG statistics were computed at the participant level to avoid treating multiple epochs from the same participant as independent observations.

Each EEG recording was preprocessed using a common signal-processing pipeline, including filtering, artefact control and segmentation into short windows. Window-level features were used only to estimate stable participant-level descriptors. For each window, relative band power was computed as

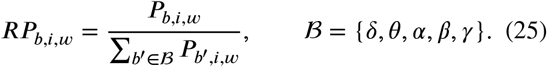

The window-level spectral state was

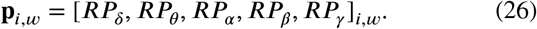

Participant-level features included theta-to-alpha ratio, individual alpha frequency, spectral entropy, Lempel-Ziv complexity and spectral flexibility. Spectral entropy was computed as

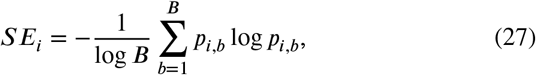

and spectral flexibility was defined as the average displacement between consecutive spectral states:

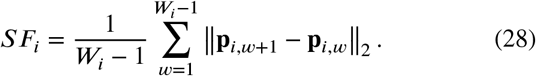

Features were normalised using the cognitively normal group:

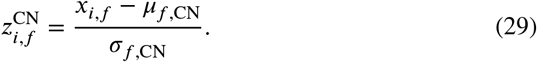

The Neurodynamic Rigidity Index was constructed as a signcorrected unweighted sum:

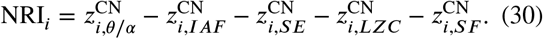

Each term was oriented so that a larger contribution indicated greater rigidity. An unweighted sum was used to avoid training a cohort-specific weighting scheme on a small EEG sample. Higher NRI values therefore indicate stronger spectral slowing, lower alpha frequency, lower entropy, lower complexity and reduced spectral flexibility. In the notation of HCCB, NRI was treated as an inverse empirical proxy for neurodynamic flexibility:

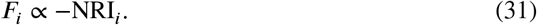

Group differences in resting NRI were tested with

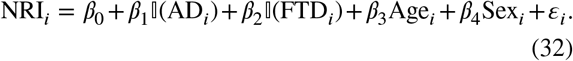

The relationship between rigidity and global cognition was evaluated by

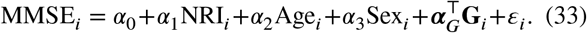

For ds006036, baseline and stimulation segments were represented using the same feature space. Stimulation adaptability was defined as the CN-referenced distance between stimulation and baseline states:

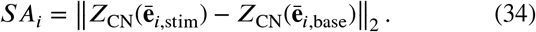

This stimulation analysis was treated as exploratory because the primary EEG test concerned resting neurodynamic rigidity rather than direct measurement of memory consolidation.

## 4. Experimental setup

### 4.1. Datasets and Cohort Definition

The experiments used three data sources corresponding to the three empirical levels of the proposed framework: clinical memory behaviour, structural MRI and electrophysiological dynamics. ADNI was used for the clinical and MRI analyses. Diagnostic records, demographic variables, APOE4 genotype, neuropsychological measures and processed MRI features were harmonised at the participant level using the ADNI participant identifier. The primary ADNI analysis used a 36-month conversion window and included four groups: cognitively normal participants, MCI nonconverters, MCI converters and AD patients. MCI converters were defined as baseline MCI participants who converted to AD within 36 months. MCI nonconverters were defined as baseline MCI participants who did not convert within the same window, including participants who converted after 36 months. Participants with insufficient follow-up and no observed conversion were excluded from the fixed-window conversion analysis. The resulting ADNI cohort comprised 1570 cognitively normal participants, 534 MCI nonconverters, 272 MCI converters and 556 AD patients.

Logical Memory was used as the primary task for bottleneck phenotyping because it provides immediate and delayed recall measures within the same assessment. Logical Memory I was used as the immediate encoding measure, and Logical Memory II was used as the delayed retention measure. RAVLT was used as a secondary memory task, with trial 5 used as the immediate reference and 30-minute delayed recall used as the delayed measure. Structural MRI validation used processed regional measures rather than raw MRI images. The main structural variables were intracranial volume-adjusted hippocampal volume and entorhinal cortical thickness. Hippocampal volume was divided by estimated total intracranial volume before analysis, and MRI predictors were standardised before regression modelling (Jack Jr et al., 2008).

OpenNeuro EEG data were used to test the neurodynamic assumption of HCCB. The resting-state dataset ds004504 was used as the primary EEG dataset and included 29 cognitively normal controls, 23 frontotemporal dementia patients and 36 AD patients (Schwerin et al., 2025). The photic-stimulation dataset ds006036 was used as an exploratory stimulation-adaptability dataset with the same diagnostic grouping. The EEG analysis was conducted at the participant level. Multiple EEG windows were used to estimate spectral and complexity features, but statistical inference was performed only on participant-level summaries to avoid epoch-level pseudo-replication.

### 4.2. Implementation and Evaluation Protocol

All ADNI analyses were performed on a participant-level baseline table. The primary outcome was 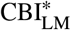, the cognitively normal reference-normalised residual Consolidation Bottleneck Index derived from Logical Memory. The cognitively normal group was used to train the reference model predicting delayed recall from immediate recall, age, sex and education. This model was then applied to all participants to estimate the expected delayed recall score. The difference between expected and observed delayed recall, normalised by the residual standard deviation in the cognitively normal group, was used as 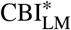. Group comparisons were performed using robust linear^L^m^M^odels with HC3 standard errors. The main covariates were age, sex, education and APOE4 dose. For MRI mechanism analysis, hippocampal volume and entorhinal thickness were tested in separate and joint models. For conversion analysis, staged logistic models were evaluated for 36-month MCI-to-AD conversion using AUC. The base model included demographic and genetic variables, the clinical model added MMSE, ADNI-MEM and CDR-SB, the structural model added hippocampal volume, and the full model added 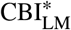.

For EEG, each participant wa^L^s^M^represented by a set of spectral and dynamical features. Resting EEG features included relative band power, theta-to-alpha ratio, individual alpha frequency, spectral entropy, Lempel-Ziv complexity and spectral flexibility. All features were normalised using the cognitively normal group as reference. The Neurodynamic Rigidity Index was computed as a sign-corrected sum in which larger values reflected spectral slowing, reduced alpha frequency, lower entropy, lower complexity and reduced spectral flexibility. Group differences in NRI were tested with linear models adjusted for age and sex. The association between NRI and global cognition was evaluated using MMSE. For ds006036, stimulation adaptability was computed as the cognitively normal referenced distance between stimulation and baseline EEG feature states. This stimulation analysis was treated as exploratory, while resting NRI in ds004504 was considered the primary EEG test.

## 5. Results and discussion

### 5.1. HCCB dynamics predict consolidation bottlenecks

The HCCB model was designed to separate two mechanisms that are usually conflated in clinical memory scores: weak initial encoding and failure to stabilise information after encoding. In the model, newly presented information first forms a rapidly decaying hippocampal trace, while a slower cortical trace accumulates through consolidation. Figure 2a shows that increasing disease load weakened the hippocampal trace and reduced the build-up of the cortical trace, producing a growing gap between immediate availability and later retention. This gap was quantified as a Consolidation Bottleneck Index, defined as the loss of delayed recall relative to immediate recall. Figure 2b shows that the short clinical-delay bottleneck remained small across simulated stages, whereas the longer consolidation-window bottleneck increased from cognitively normal ageing to stable MCI, MCI conversion and AD (Jutten et al., 2025). This pattern formalises the central idea of the framework: new-memory failure in AD is not only a reduction in what is initially encoded, but also a failure to preserve encoded information over time.

**Figure 2:**
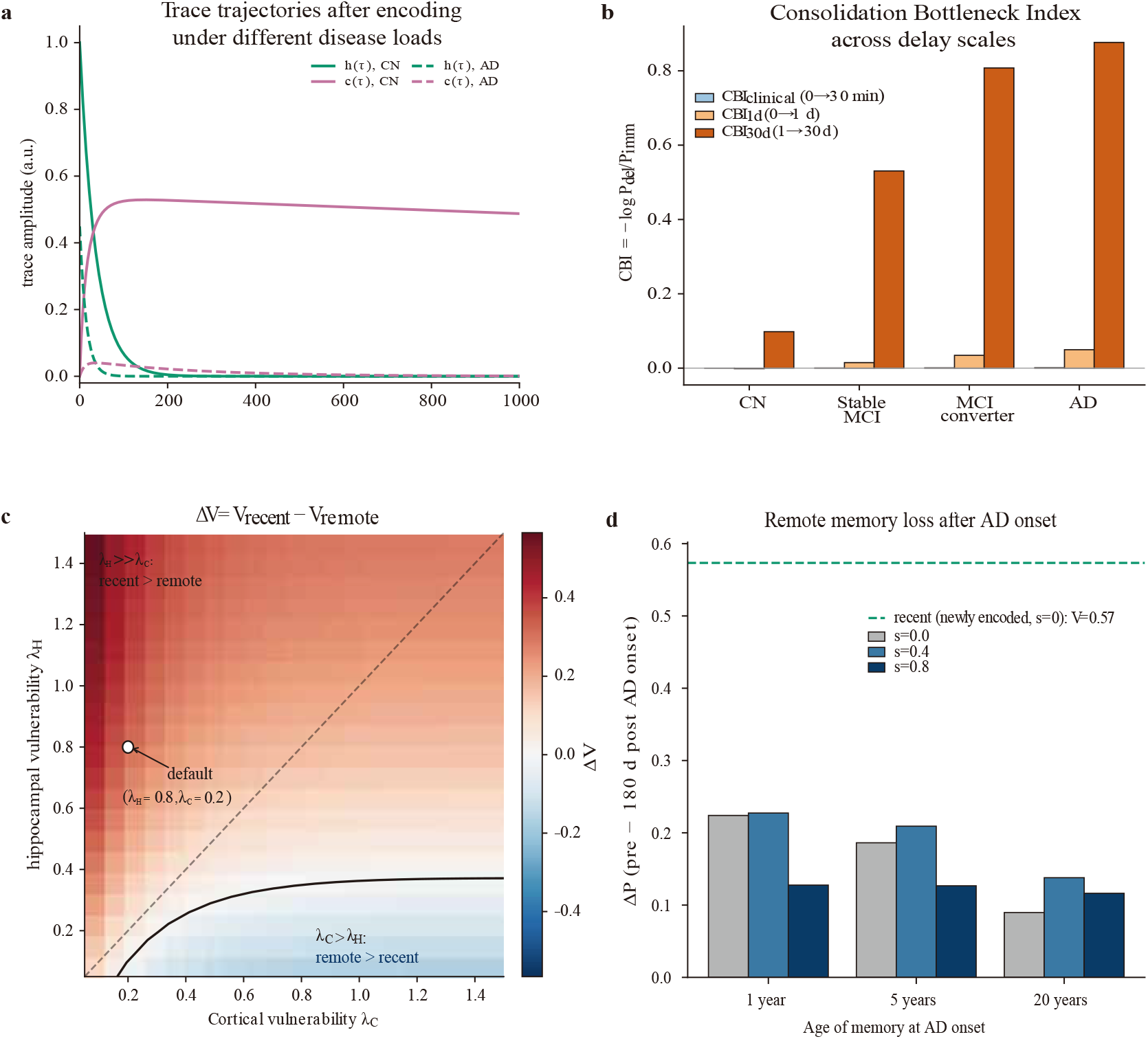
HCCB dynamics generate consolidation bottlenecks and recent-memory vulnerability. a, Simulated hippocampal and cortical memory traces after encoding under cognitively normal and AD-like disease loads. The hippocampal trace decays rapidly, whereas the cortical trace accumulates slowly through consolidation; AD-like load weakens both early trace stability and later cortical stabilisation. b, Consolidation Bottleneck Index across delay scales and simulated clinical stages. Longer consolidation-window bottlenecks increase from cognitively normal ageing to stable MCI, MCI conversion and AD, whereas the short clinical-delay bottleneck remains comparatively small. c, Parameter-space analysis of recent versus remote vulnerability. When hippocampal vulnerability exceeds cortical vulnerability, recent memories are predicted to be more disease-sensitive than remote memories; the opposite regime appears when cortical vulnerability dominates. The default AD-like parameter setting lies in the recent-vulnerability region. d, Two-phase simulation of remote memory loss after AD onset. Memories consolidated before disease onset show smaller post-onset decline than newly encoded recent memories, with stronger schema support further reducing vulnerability.

The model further generated a parameter-level prediction for temporal memory vulnerability. Figure 2c shows that when hippocampal vulnerability exceeded cortical vulnerability, the system entered a regime in which recent memories were more disease-sensitive than remote memories. Conversely, when cortical vulnerability dominated, the model predicted the opposite pattern, indicating that the temporal gradient was not imposed by assumption but emerged from the relative vulnerability of hippocampal and cortical components. A two-phase simulation then tested the effect of disease onset on memories that had already undergone prior consolidation. Figure 2d shows that newly encoded recent memories had the largest post-onset loss, whereas memories formed one, five or twenty years before disease onset showed smaller declines, especially when schema support was stronger. Together, these simulations define the empirical predictions tested in subsequent analyses: a residual delayed-retention bottleneck should be detectable in clinical memory data, should be strongest in individuals approaching AD conversion, and should be linked to hippocampal system integrity.

### 5.2. ADNI reveals an AD-like bottleneck in MCI converters

We next tested whether the residual consolidation bottleneck predicted by HCCB could be detected in ADNI clinical memory data. The cohort included cognitively normal participants, MCI nonconverters, MCI converters and AD patients, with the expected clinical gradient in MMSE, CDRSB, ADNI-MEM (Wilkosz et al., 2010), Logical Memory immediate recall and Logical Memory delayed recall as shown in Table 1. To isolate post-encoding stabilization failure from poor initial learning, 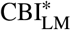 was defined as delayed Logical Memory recall lower than expected from immediate recall and demographic factors in a cognitively normal reference model. Figure 3a shows that 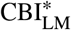 increased across the AD clinical spectrum, from CN to MCI nonconverters, MCI converters and AD. The strongest transition-stage pattern was observed in MCI converters, whose 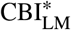 was higher than that of MCI nonconverters and statistica^L^ll^M^y comparable to AD, rather than significantly exceeding AD. This pattern supports the interpretation that MCI converters already express an AD-like stabilization-failure phenotype before clinical conversion.

**Table 1.** Demographic, clinical, cognitive and MRI characteristics of the ADNI cohort.

| Variable | CN | MCI nonconv. | MCI conv. | AD | Test |
| --- | --- | --- | --- | --- | --- |
| *n* | 1570 | 534 | 272 | 556 | – |
| Age, yr | 70.6 (7.2) | 72.1 (7.6) | 73.8 (7.2) | 74.9 (8.0) | $F = 54.0, p < 0.001$ |
| Female, n/N (%) | 967/1570 (61.6) | 217/534 (40.6) | 109/272 (40.1) | 243/556 (43.7) | $\chi^2 = 116.2, p < 0.001$ |
| Education, yr | 16.5 (2.5) | 16.1 (2.7) | 16.0 (2.7) | 15.3 (2.9) | $F = 30.3, p < 0.001$ |
| APOE4 dose | 0.37 (0.56) | 0.48 (0.63) | 0.82 (0.67) | 0.87 (0.72) | $F = 91.5, p < 0.001$ |
| MMSE | 29.00 (1.21) | 27.99 (1.72) | 26.74 (1.77) | 22.99 (2.80) | $F = 1616.8, p < 0.001$ |
| CDR-SB | 0.07 (0.24) | 1.28 (0.78) | 2.07 (1.00) | 4.49 (1.85) | $F = 3059.9, p < 0.001$ |
| ADNI-MEM | 1.201 (0.692) | 0.480 (0.656) | -0.385 (0.559) | -0.993 (0.642) | $F = 1192.0, p < 0.001$ |
| LM I immediate | 13.73 (3.47) | 9.57 (3.38) | 6.62 (3.47) | 4.20 (3.09) | $F = 1246.9, p < 0.001$ |
| LM II delayed | 12.67 (3.53) | 6.96 (3.25) | 3.52 (3.12) | 1.64 (2.41) | $F = 1922.5, p < 0.001$ |
| $CBI^*_{LM}$ | -0.000 (1.000) | 1.138 (1.115) | 1.639 (1.249) | 1.546 (1.121) | $F = 435.7, p < 0.001$ |
| $CBI_{LM}$ | 0.085 (0.172) | 0.397 (0.502) | 0.813 (0.815) | 1.037 (0.951) | $F = 483.2, p < 0.001$ |
| RAVLT trial 5 | 11.63 (2.47) | 9.41 (2.79) | 6.71 (2.30) | 5.24 (2.04) | $F = 887.0, p < 0.001$ |
| RAVLT delayed | 7.91 (3.98) | 4.96 (3.92) | 1.68 (2.42) | 0.72 (1.60) | $F = 593.6, p < 0.001$ |
| $CBI_{RAVLT-T5}$ | 0.537 (0.662) | 0.935 (0.851) | 1.682 (0.900) | 1.893 (0.763) | $F = 444.8, p < 0.001$ |
| Hippocampus/eTIV | 0.0050 (0.0006) | 0.0047 (0.0007) | 0.0041 (0.0006) | 0.0040 (0.0006) | $F = 349.2, p < 0.001$ |
| Entorhinal, mm | 3.423 (0.278) | 3.301 (0.409) | 2.953 (0.445) | 2.777 (0.471) | $F = 325.6, p < 0.001$ |

Values are mean (SD) unless otherwise stated. CN, cognitively normal; MCI, mild cognitive impairment; AD, Alzheimer's disease; LM, Logical Memory; MMSE, Mini-Mental State Examination; CDR-SB, Clinical Dementia Rating Sum of Boxes; RAVLT, Rey Auditory Verbal Learning Test; eTIV, estimated total intracranial volume; CBI, Consolidation Bottleneck Index.

**Figure 3:**
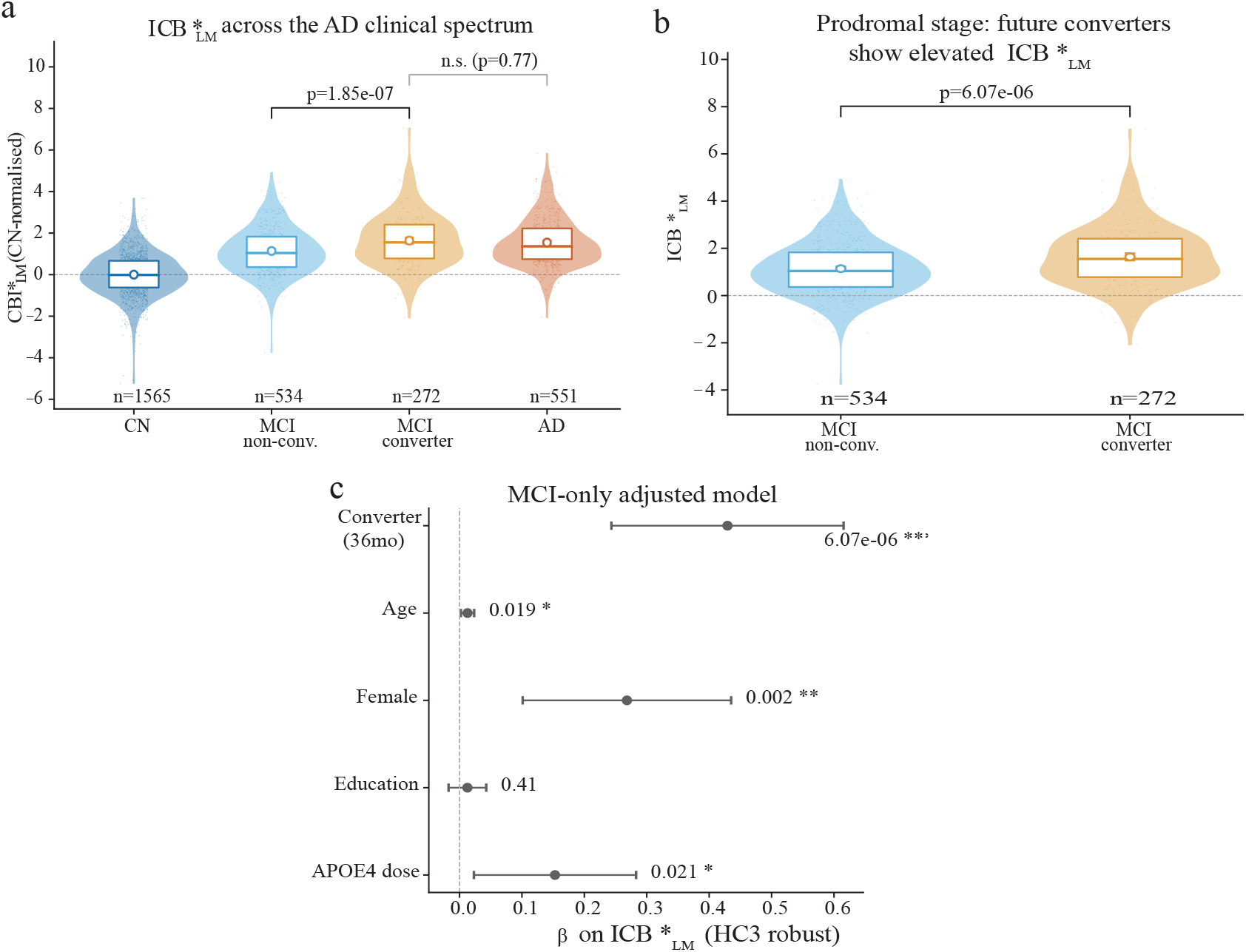
ADNI reveals an AD-like residual consolidation bottleneck in MCI converters. a, Distribution of 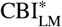 across the AD clinical spectrum. 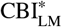 was lowest in cognitively normal participants, elevated in MCI nonconverters, further increased in MCI converters and remained high in AD. MCI converters showed higher 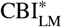 than MCI nonconverters, while their difference from AD was not significant. b, MCI-only comparison showing elevated 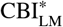 in future 36-month converters relative to nonconverters. c, MCI-only adjusted model for 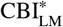. Converter status remained as sociated with higher 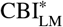 after adjustment for age, sex, education and APOE4 dose, supporting a transition-stage residual stabilization-failure phenotype.

We then focused on baseline MCI participants to test whether this residual bottleneck distinguished future converters from nonconverters within the same clinical stage. Figure 3b shows that MCI converters had higher 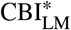 than MCI nonconverters. The MCI-only adjusted model in Figure 3c confirmed that converter status remained associated with higher 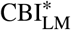 after accounting for age, sex, education and APOE4 dose, with *β* = 0.429 and *p* < 0.001. These results indicate that the HCCB-derived bottleneck is not merely a reflection of broad disease-stage differences between CN and AD (Choe et al., 2020). Instead, it captures a specific residual memory-stabilization deficit that is already detectable among MCI individuals who later progress to AD.

### 5.3. Consolidation bottleneck links to hippocampal atrophy

The HCCB framework predicts that a residual stabilization deficit should not behave as an arbitrary cognitive score, but should be anchored to the hippocampal system that supports early memory formation. Figure 4a shows that higher 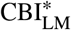 was associated with lower ICV-adjusted hippocampal volume after adjustment for age, sex, education, APOE4 and diagnosis. The partial association was negative, with = 0.224 per SD, *p* = 3.22∝10^−12^ and n = 2007. The primary MRI models in Table 2 further supported this relationship. In the full sample, standardized hippocampal volume was associated with lower 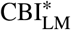 in the hippocampus-only model, and the association remained significant in the joint model including entorhinal thickness. The same direction was observed in the MCI-only analysis, where hippocampal volume showed an even larger standardized association with 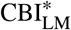. These results indicate that the residual bottleneck captured by 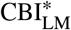 is structurally linked to hippocampal degeneration rather than reflecting only broad diagnostic status (Dodge et al., 2014). We next examined whether 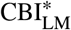 should be interpreted as a prognostic replacement for established clinical and structural markers. Figure 4b shows that CBI^∗^ alone yielded modest discrimination for 36-month MCIto-AD conversion, with an AUC of 0.617. Adding clinical variables produced a much larger gain, and the structural model reached an AUC of 0.885. Adding 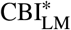 to the clinical and structural model increased AUC only from 0.885 to 0.887, with a non-significant DeLong comparison against the structural model. This pattern clarifies the role of 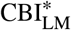 in the study: it is not intended to replace established prognostic markers such as ADNI-MEM, CDR-SB or hippocampal volume (Li et al., 2017). Instead, it provides a mechanistically interpretable phenotype of post-encoding stabilization failure that is elevated in MCI converters and biologically aligned with hippocampal atrophy.

**Table 2.** Primary ADNI statistical models for residual consolidation bottleneck and 36-month MCI conversion.

| A. Adjusted group contrasts for CBI*<sub>LM</sub> across the AD clinical spectrum |  |  |  |  |  |
| --- | --- | --- | --- | --- | --- |
| Effect / contrast | $\beta$ | 95% CI | $p$ | $n$ | |
| MCI nonconverter vs CN | 1.197 | 1.083 to 1.310 | $p < 0.001$ | 2397 | |
| MCI converter vs CN | 1.673 | 1.507 to 1.839 | $p < 0.001$ | 2397 | |
| AD vs CN | 1.646 | 1.516 to 1.776 | $p < 0.001$ | 2397 | |
| MCI converter vs MCI nonconverter | 0.476 | 0.297 to 0.655 | $p < 0.001$ | 2397 | |
| AD vs MCI nonconverter | 0.449 | 0.303 to 0.595 | $p < 0.001$ | 2397 | |
| MCI converter vs AD | 0.027 | -0.153 to 0.207 | 0.769 | 2397 |  |

| B. MCI-only adjusted model for CBI*<sub>LM</sub> |  |  |  |  |  |
| --- | --- | --- | --- | --- | --- |
| Predictor | $\beta$ | 95% CI | $p$ | $n$ | |
| MCI converter vs MCI nonconverter | 0.429 | 0.243 to 0.615 | $p < 0.001$ | 805 | |
| Age | 0.013 | 0.002 to 0.024 | 0.0187 | 805 |  |
| Female sex | 0.268 | 0.101 to 0.435 | 0.00164 | 805 |  |
| Education | 0.013 | -0.018 to 0.043 | 0.413 | 805 |  |
| APOE4 dose | 0.153 | 0.023 to 0.283 | 0.0211 | 805 |  |

| C. MRI mechanism models for CBI*<sub>LM</sub> |  |  |  |  |  |
| --- | --- | --- | --- | --- | --- |
| Model | Predictor | $\beta$ | 95% CI | $p$ | $n$ |
| Hippocampus-only, full sample | $z$ hippocampal volume/eTIV | -0.235 | -0.293 to -0.177 | $p < 0.001$ | 2300 |
| Entorhinal-only, full sample | $z$ entorhinal thickness | -0.132 | -0.190 to -0.074 | $p < 0.001$ | 2300 |
| Joint MRI model, full sample | $z$ hippocampal volume/eTIV | -0.212 | -0.273 to -0.150 | $p < 0.001$ | 2300 |
| Joint MRI model, full sample | $z$ entorhinal thickness | -0.055 | -0.115 to 0.006 | 0.0776 | 2300 |
| Hippocampus-only, MCI-only | $z$ hippocampal volume/eTIV | -0.352 | -0.458 to -0.247 | $p < 0.001$ | 781 |
| Joint MRI model, MCI-only | $z$ hippocampal volume/eTIV | -0.352 | -0.461 to -0.243 | $p < 0.001$ | 781 |
| Joint MRI model, MCI-only | $z$ entorhinal thickness | -0.001 | -0.105 to 0.103 | 0.983 | 781 |

| D. Staged model comparison for 36-month MCI-to-AD conversion |  |  |  |  |
| --- | --- | --- | --- | --- |
| Model | AUC | ΔAUC vs Base | ΔAUC vs previous | |
| CBI*<sub>LM</sub> only | 0.617 | — | — |  |
| Base model | 0.674 | 0.000 | 0.057 |  |
| Clinical model | 0.885 | 0.211 | 0.211 |  |
| Structural model | 0.885 | 0.211 | 0.000 |  |
| Full model + CBI*<sub>LM</sub> | 0.887 | 0.213 | 0.002 |  |
| Full vs Structural, DeLong $p$ | — | 0.002 | 0.415 | |

CBI\*<sub>LM</sub> denotes the cognitively normal reference-normalised residual Consolidation Bottleneck Index derived from Logical Memory delayed recall. Models in panels A and B used HC3 robust standard errors and adjusted for age, sex, education and APOE4 dose. MRI predictors in panel C were $z$ -standardised; eTIV denotes estimated total intracranial volume. The staged conversion analysis in panel D evaluated 36-month MCI-to-AD conversion. The base model included age, sex, education and APOE4 dose; the clinical model added MMSE, ADNI-MEM and CDR-SB; the structural model added ICV-adjusted hippocampal volume; the full model added CBI\*<sub>LM</sub>.

**Figure 4:**
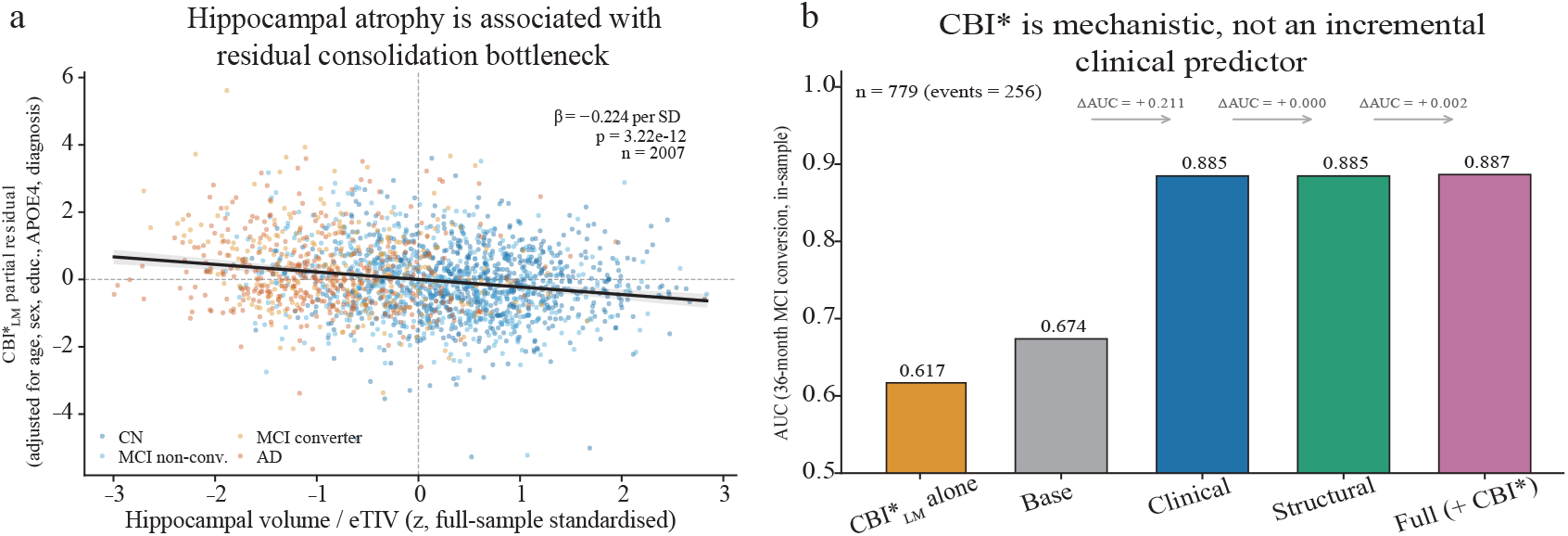
Structural anchoring and prognostic positioning of CBI*. a, Partial association between ICV-adjusted hippocampal volume and 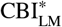 after adjustment for age, sex, education, APOE4 and diagnosis. Lower hippocampal volume was associated with higher residual consolidation bottleneck, supporting the structural relevance of 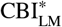. Points are coloured by diagnostic group. b, Staged model comparison for 36-month MCI-to-AD conversion. 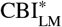 alone showed modest discrimination. Clinical and structural predictors explained most of the prognostic information, and adding 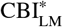 to the full model produced only a small, non-significant AUC gain. This supports the interpretation of 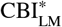 as a mechanistic phenotype rather than a standalone prognostic replacement.

### 5.4. OpenNeuro EEG supports reduced neurodynamic flexibility in AD

We finally tested the electrophysiological component of HCCB using independent OpenNeuro EEG datasets. This analysis was not intended to directly measure memory consolidation, but to test whether AD is accompanied by the reduced neurodynamic flexibility assumed by the model. In the resting-state EEG dataset ds004504, we summarised spectral slowing, alpha peak reduction, entropy loss, complexity reduction and reduced spectral flexibility into a cognitively normal referenced Neurodynamic Rigidity Index. Figure 5a shows that resting NRI was markedly elevated in AD compared with cognitively normal controls, with AD versus CN *p* < 0.001. Figure 5b decomposes this increase and shows that the group-level rigidity signal was primarily driven by increased theta-to-alpha ratio and reduced alpha frequency, with additional contributions from reduced entropy, complexity and flexibility. This pattern supports the model assumption that AD is associated with a less flexible and more rigid dynamical regime. We next examined whether this rigidity was clinically meaningful and whether it extended to stimulus-driven adaptation. Figure 5c shows that higher resting NRI was associated with lower MMSE across participants, with *r* = −0.44, *p* < 0.001 and *n* = 88, indicating that neurodynamic rigidity tracked global cognitive impairment. In the photic-stimulation dataset ds006036, stimulation adaptability was numerically altered across groups, but the AD versus CN difference did not reach significance as shown in Figure 5d, with p = 0.127. We therefore interpret the EEG results conservatively: resting EEG provides independent support for reduced neurodynamic flexibility in AD (Yang et al., 2013), while stimulation adaptability remains exploratory. Together with the ADNI findings, these results place HCCB across behavioural, structural and electrophysiological levels: ADNI identified a residual stabilization bottleneck, MRI linked it to hippocampal degeneration and EEG showed an AD-related rigid dynamical background consistent with impaired stabilization of newly encoded information.

**Figure 5:**
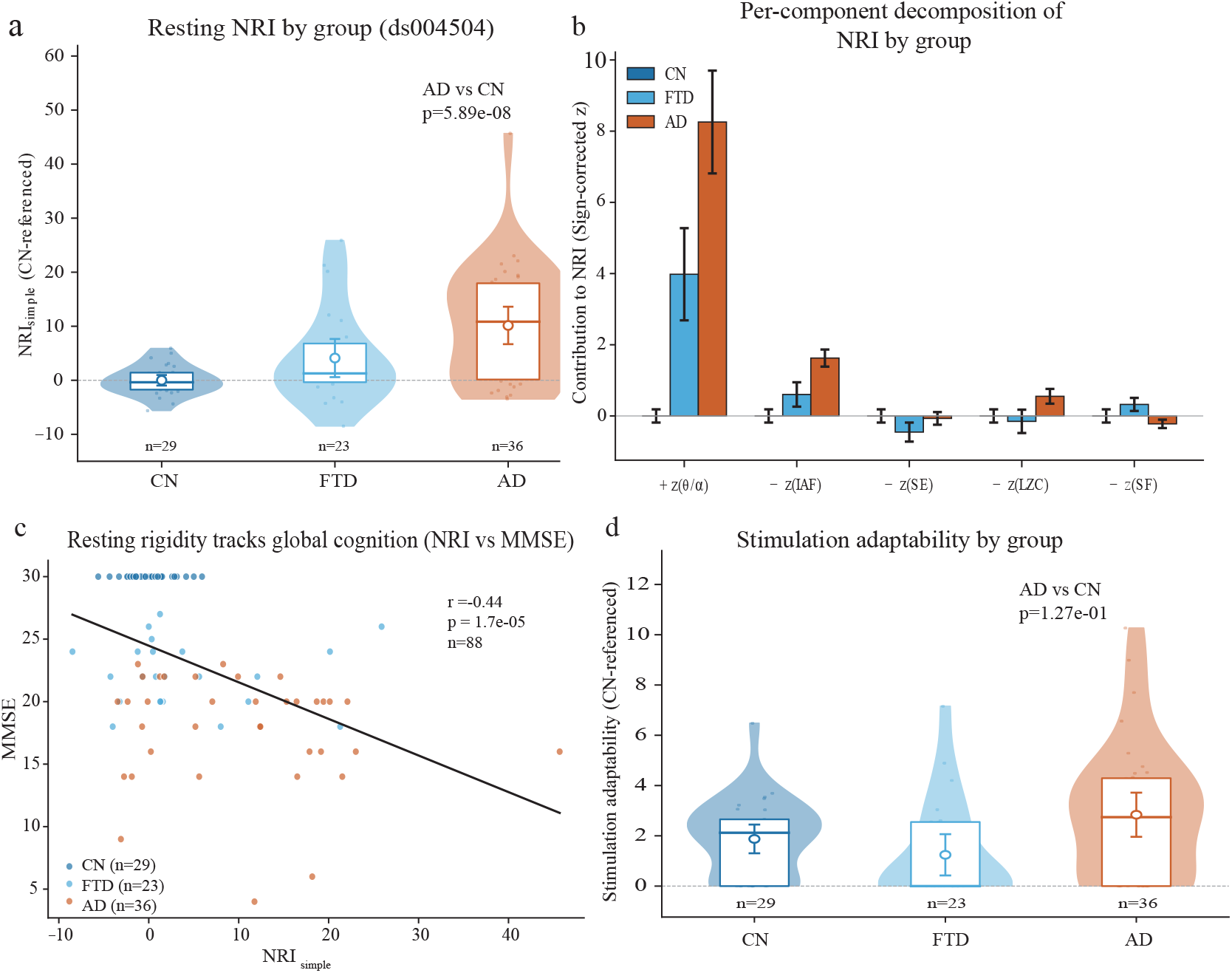
OpenNeuro EEG reveals increased neurodynamic rigidity in AD. a, Resting-state Neurodynamic Rigidity Index in ds004504 across cognitively normal controls, frontotemporal dementia and AD. AD showed higher NRI than cognitively normal controls, indicating reduced neurodynamic flexibility. b, Sign-corrected decomposition of NRI components. Increased theta-toalpha ratio and reduced individual alpha frequency contributed most strongly to AD-related rigidity, with additional contributions from spectral entropy, Lempel-Ziv complexity and spectral flexibility. c, Association between resting NRI and MMSE across all participants. Higher NRI was associated with lower global cognitive performance. d, Stimulation adaptability in the photicstimulation dataset ds006036. The AD versus CN difference was not significant and was treated as exploratory rather than as primary evidence.

## Data Availability

The ADNI data used in this study are available through the Alzheimer's Disease Neuroimaging Initiative data portal and require a formal data access application and approval in accordance with ADNI data use policies. The OpenNeuro EEG datasets analysed in this study, including ds004504 and ds006036, are publicly available through the OpenNeuro platform. No raw ADNI data are redistributed with this article. The code used for data processing, HCCB simulation, ADNI analysis, EEG feature extraction and figure generation is available at: https://github.com/Chriszhangmw/ad-computation.

## 6. Conclusions and future work

This paper presented HCCB, a computational framework for interpreting new-memory failure in Alzheimer’s disease by separating poor initial encoding from failed postencoding stabilisation. The resulting residual bottleneck phenotype, CBI^∗^, converts this distinction into a measurable clinical index derived from routine immediate and delayed memory testing. Experiments on ADNI showed that MCI converters exhibited an Alzheimer-like elevation of CBI^∗^ before clinical conversion, and that higher CBI^∗^ was associated with hippocampal atrophy. These results indicate that the transition toward Alzheimer’s disease involves a specific stabilisation failure that is not captured by delayed recall alone. OpenNeuro EEG analyses further supported the dynamical assumption of the framework by showing increased neurodynamic rigidity in Alzheimer’s disease. Overall, the study provides a model-driven computational phenotype for characterising how newly encoded information fails to become stable during Alzheimer’s disease progression.

Future work should extend this framework to datasets that contain memory testing, MRI and EEG from the same participants, enabling direct subject-level testing of the link between CBI^∗^ and neurodynamic rigidity. The present study is limited by the use of independent ADNI and OpenNeuro cohorts, and by the absence of dense memory measurements across minutes, days and weeks. Cohorts with repeated consolidation-sensitive memory tasks, sleep or task EEG, and longitudinal imaging would allow the full HCCB dynamics to be fitted more directly and may support personalised modelling of encoding and stabilisation failure.

## CRediT authorship contribution statement

**Meiwei Zhang:** Conceptualization, Methodology, Software, Writing original draft. **Yuwei Pan:** Data Curation. **Lihua Chen:** Resources.

## Declaration of generative AI in scientific writing

The authors retain full responsibility and accountability for the entirety of this work, affirming the absence of generative AI in its conception and development.

## Ethical, Review board approval

This study used only secondary, de-identified data obtained from publicly available or controlled-access research repositories. The present analysis did not involve the recruitment of new human participants, any new clinical intervention, clinical decision-making, or the use of identifiable personal information. Therefore, no additional institutional ethical approval was required for this secondary analysis. ADNI data were accessed through the Alzheimers Disease Neuroimaging Initiative data access process and used in accordance with ADNI data use policies. The OpenNeuro EEG datasets were publicly available and analysed according to their data use terms. All original data collection procedures were conducted by the respective data providers under their approved ethical protocols and informed consent procedures. This study did not involve any animal experiments.

